# A secondary data analysis investigating the link between weather elements and incident presentation for mental disorders at a Ugandan tertiary psychiatric hospital

**DOI:** 10.1101/2025.01.31.25321476

**Authors:** Emmanuel Kiiza Mwesiga, Ian Munabi, Andrew Sentoogo Ssemata, Allan Kalungi, Sophia Balinga, Blessed Tabitha Aujo, Bryan Byamah Mutamba, Martha Bbosa, Robert Kalyesubula, Fred Babweetera, Eugene Kinyanda, Wilber Ssembajjwe

## Abstract

The link between weather elements and mental disorders is often described in high-income countries, with hardly any data from low-and middle-income countries where the resources to cope with the negative mental health impacts of climate change are extremely constrained. In this paper, we examined the association between weather elements and the incident presentation with a mental disorder at Butabika National Referral Mental Hospital. We used secondary data from two datasets: i) a mental health data set collated from all patients presenting at Butabika National Referral Mental Hospital in 2019; and ii) a climatic dataset for the geographic location of Butabika National Referral Mental Hospital for the same year (2019). The mental health data set included socio-demographic variables and mental disorder diagnoses, while the climatic data set included data on, atmospheric pressure (hPa), rainfall (mm), sunshine (hours/month), humidity (%), temperature (°C) and wind speed (m/s). We performed descriptive statistical analyses to summarize the frequency of mental disorder diagnoses and the monthly averages of weather variables. We then undertook correlation and multiple logistic regression analyses to investigate the associations between specific weather elements and the incident presentation of different mental disorders. In the mental health data, we had 2,827 participants, males were 56.1%(n=1,584), and the median age was 29 years (IQR 23-38). Psychotic disorders were the most common diagnosis at 43.8% (n=1,239). Overall, various weather elements correlated at different strengths with incident presentation of various mental disorders, particularly humidity and rainfall. On controlling for age and sex in the multiple regression models, the strongest associations were between heat elements and incident presentation for psychotic disorders [AOR1.12, 95%CI (1.04; 1.27) p<0.001]. No associations were demonstrated between weather elements and incident presentation for neurocognitive and neurodevelopmental disorders. These preliminary findings point to a possible relationship between incident presentation of mental disorders to a tertiary psychiatric hospital in Uganda and various weather elements. There is need for longitudinal studies to confirm these associations and to explore underlying social and biological mechanisms.

## Introduction

The intricate relationship between weather elements and mental health has garnered increasing attention in recent years, particularly as global concerns about climate change continue to rise (1, 2). Evidence suggests that weather elements, such as temperature, humidity, atmospheric pressure, and rainfall, can significantly influence mental health outcomes. These influences can manifest in various ways, including the onset of mental health conditions (3), worsening of existing symptoms (4), and even triggering acute mental health crises (5). For instance, extreme temperatures have been associated with increased rates of hospital admissions for mood disorders (6, 7). At the same time, changes in atmospheric pressure have been linked to alterations in the severity of depressive and anxiety symptoms (8, 9). Despite increasing evidence of such relationships in high-income, temperate regions, research remains sparse in many low- and middle-income countries, particularly in sub-Saharan Africa. In these regions, there is increasing evidence that the effects of climate change are having a more significant impact. For example, in East Africa, Uganda experiences a tropical climate characterized by alternating wet and dry seasons (10), and incident presentations for various mental disorders vary across the year and seasons.

Further work is required to examine the link between weather elements and incident presentation with a mental disorder in low-resource settings like Uganda. First, it is essential to investigate a link between weather elements, including temperature, humidity, atmospheric pressure, and rainfall and the incident presentation for mental disorders. Further, it is unclear if the weather elements have similar strengths of association with the different diagnoses, including mood, anxiety, psychotic, neurocognitive, neurodevelopmental, and substance use disorders. Lastly, the role of sociodemographic factors, including age and sex, in the link between weather elements and incident presentation for a mental disorder requires further review.

By analyzing historical weather data alongside hospital records, this study aims to investigate the associations between specific weather elements and the incident presentation of mental disorders in Uganda. Advancements in precision medicine require the integration of diverse data points, including environmental factors. Although considerable emphasis has been placed on psychological factors like trauma, weather elements remain underexplored despite their pervasive and mediating influence on factors such as diet and employment. factors such as diet and employment. These findings will contribute to the limited body of knowledge on environmental determinants of mental health in low-resource settings and inform public health strategies and interventions, particularly in the context of climate change and its impact on population health.

## Materials and Methods

We undertook secondary data analyses using a mental health dataset from Butabika National Referral Mental Hospital in Uganda and climatic data from Entebbe Meteorological Centre. Butabika National Referral Mental Hospital is the largest psychiatric facility in Uganda, which serves as the primary referral center for individuals with severe mental disorders across the country. Patients reach the hospital via referrals from regional hospitals, healthcare centers, and sometimes arrive directly with the support of their families or community health workers. The hospital predominantly serves patients presenting with a wide range of psychiatric conditions, including schizophrenia, mood disorders, substance use disorders, and other severe mental health issues, as well as general medical conditions. Data at this hospital are routinely captured during patient intake and follow-up, with trained healthcare workers systematically recording clinical information, demographics, and presenting symptoms into a central database. A description of both datasets is given below:

### Data on incident presentation with a mental disorder

This was a retrospective analysis of chart records on Friday 17 May 2024 to identify patients presenting to the hospital for the first time (11). Ethical approval for the study was obtained from the School of Medicine Research and Ethics Committee (SOMREC) of Makerere University and Uganda National Council for Science and Technology (UNCST) (#REC REF 2017-086). We also received institutional approval from the hospital to conduct the study.

Because this was a retrospective chart review of file records, we requested a waiver for participant consent. The study included all patients presenting to Butabika National Referral Mental Hospital for the first time between January 1st and December 31st, 2019. within a 50km radius from Entebbe Meteorological Centre. We excluded patients presenting for the first time for non-psychiatric services such as dental services, routine HIV care, and minor surgeries such as circumcision. Hospital clinicians documented diagnoses based on the *Diagnostic and Statistical Manual of Mental Disorders 5^th^ edition (DSM-5)* (12). Data abstraction was performed on to extract sociodemographic information, including age, gender, and home district, to provide contextual insights into the patient population.

### Data on weather elements

The weather elements dataset is part of a larger dataset of nine purposively selected study districts in Uganda, whose health and weather data were available for the development of an early warning health model (https://github.com/CHAIUGA/chasa-model) and an accompanying prediction web app (https://github.com/CHAIUGA/chasa-webapp). Historical weather data was retrieved from the Uganda National Meteorological Association database as monthly averages. The weather variables in this data included atmospheric pressure, rainfall, solar radiation, humidity, temperature (maximum, minimum and mean), and wind (gusts and average wind speed). The districts included were "Nakasongola", "Entebbe", "Butambala", "Gulu", "Kampala", "Sembabule", "Soroti", "Kitgum" and "Nakaseke".

### Data selection, merging and cleaning

For this study, we extracted from the two datasets clinical records of patients presenting at Butabika National Referral Mental Hospital and climate data from the Entebbe Meteorological Centre, covering January 2019 to December 2019. Clinical data was selected for patients from the greater Kampala Metropolitan Area (GKMA), including Kampala, Wakiso and Mukono districts. The clinical dataset comprised patient records, including diagnostic information and the date of the first presentation, collected as part of routine care. The dataset focused on new mental disorder diagnoses made according to the DSM-5 criteria and included key sociodemographic variables such as age, sex, and home district. The climate dataset was retrieved from the Entebbe Meteorological Centre, which collects monthly weather data for the Greater Kampala Metropolitan Area (GKMA), where Butabika National Referral Mental Hospital is located. The weather elements included atmospheric pressure, rainfall, sunshine hours, humidity, temperature and wind speed. Data merging involved aligning the clinical and climate datasets by month, as both datasets contained information at the monthly level. Following the merging process, data cleaning was performed to ensure completeness and accuracy. Outliers were identified and checked for plausibility but retained due to their clinical and meteorological relevance. To ensure compatibility between datasets, we conducted variable harmonization, ensuring that the units of measurement for weather variables were consistent across all months.

### Data analysis

The merged dataset was analyzed to explore correlations between weather elements and mental disorder presentations. Descriptive statistics were used to summarize the frequency of mental disorder diagnoses. The monthly averages of weather variables such as atmospheric pressure, rainfall, sunshine, humidity, temperature, and wind speed were similarly summarized. Correlation coefficients calculated the relationship between weather elements and mental disorder presentations. Multiple logistic regression models were employed to assess the associations between specific weather variables and the likelihood of presentation for different mental disorder diagnoses, including psychotic disorders, mood disorders, anxiety disorders, and substance use disorders. Odds ratios (ORs), adjusted odds ratios (AORs), 95% confidence intervals (CIs), and p-values were reported for each weather element in relation to the mental disorder outcomes. Model fit was assessed using likelihood ratio tests. Statistical significance was set at p < 0.05. All analyses were conducted using STATA version 15 (13).

## Results

### Descriptive statistics

The combined dataset had 2,827 participants (see flow chart in Fig. 1), with males at 56.1% (n=1,584), and a median age of 29 years (IQR 23-38). The nature of mental disorders were: psychotic disorders 43.83% (n=1,239), substance use disorders 16.8% (n=476), mood disorders 15.5% (n=437), neurological disorders 12.4% (n=3500), neurocognitive disorders 2.4% (n=68), anxiety disorders 1.66% (n=47), and developmental disorders 0.32% (n=9). . Additionally, 7.11% (n=201) of the cases had other psychiatric diagnoses.

**Figure 1:**
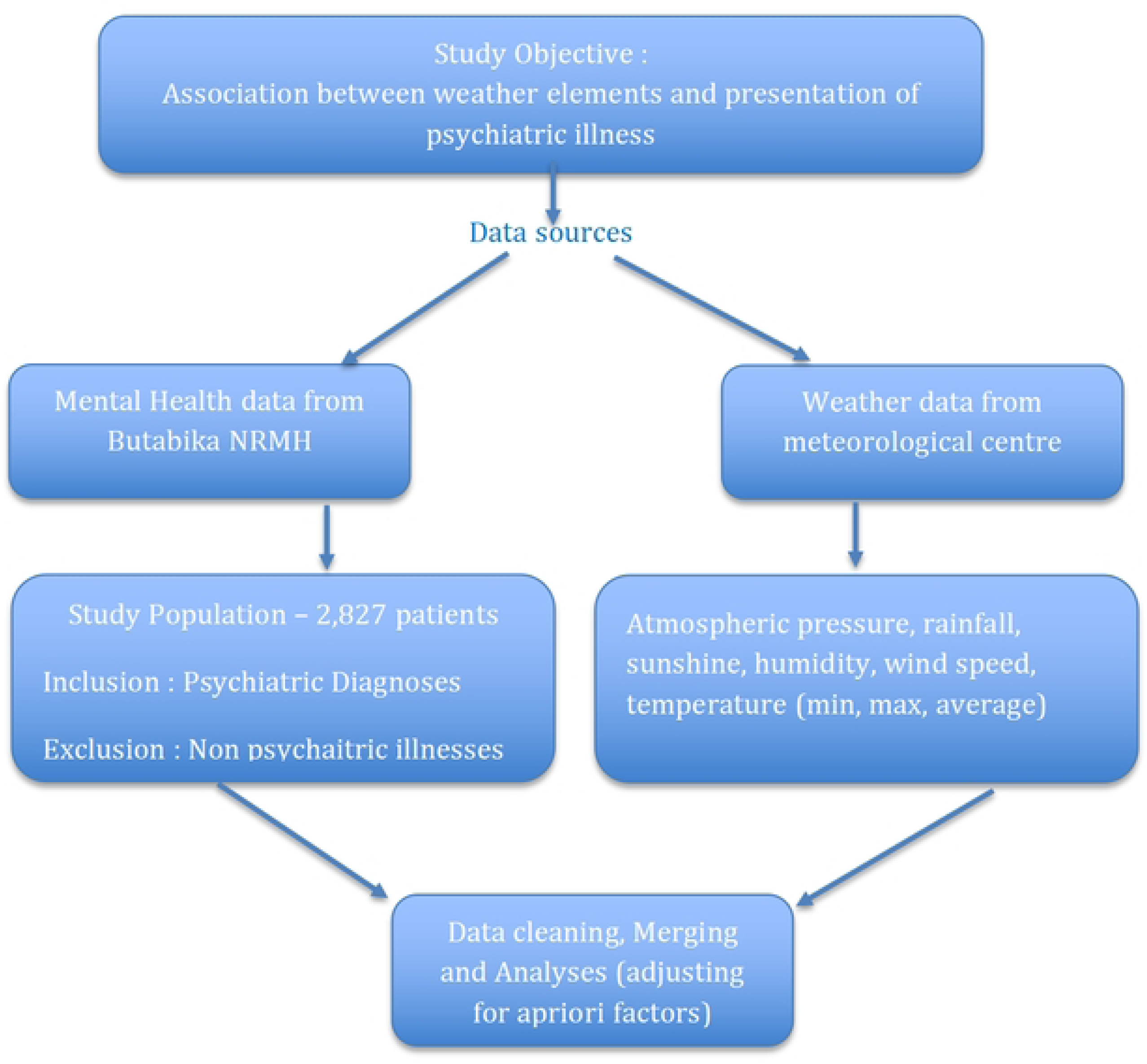
Study flowchart. Study flowchart for the selection of participants

### Climate Factors

Regarding climatic factors (Table 1), the atmospheric pressure ranged from a minimum of 88.79 hPa to a maximum of 89.17 hPa, with an average of 88.94 hPa. Rainfall varied widely, with a minimum of 33.94 mm, an average of 117.02 mm, and a peak of 173.41 mm. The sunshine duration ranged from 163.80 hours/month to 253.63 hours/month, averaging 204.45 hours/day. Humidity levels showed a narrow range, with a minimum of 63.0%, an average of 80.8%, and a maximum of 87.4%. The mean temperature recorded values from 21.48°C to 25.65°C, averaging at 22.69°C. The mean wind speed fluctuated between 0.89 m/s and 2.44 m/s, with an average of 1.17 m/s.

**Table 1:** Demographic characteristics and Summary of weather elements between Jan to Dec 2019 in Kampala.

| Factor | Level | Frequency (%) |
| --- | --- | --- |
| Age | Median (IQR) | 29 (23; 38) |
| Sex | Male | 1,584 (56.1) |
| Religion | Christian | 2,343 (82.9) |
|  | Moslem | 424 (15.0) |
|  | Non-religious / others | 59 (2.1) |

| Weather Element | Minimum | Maximum | Average |
| --- | --- | --- | --- |
| Atmospheric pressure (hPa) | 88.7922 | 89.1692 | 88.937 |
| Rain (mm) | 33.9447 | 173.413 | 117.022 |
| Sun (hours/month) | 163.802 | 253.625 | 204.45 |
| Humidity (%) | 0.630029 | 0.873911 | 0.808034 |
| Mean Temperature (°C) | 21.4783 | 25.6453 | 22.693 |
| Mean Wind Speed (m/s) | 0.893892 | 2.44071 | 1.1696 |
| Heat (max temp °C) | 29.2 | 34.9 | 31.7 |

| Mental disorder diagnoses |  |
| --- | --- |
| Diagnosis | Freq (%) |
| Psychotic disorders | 1,239 (43.8) |
| Substance Use disorders | 476 (16.8) |
| Mood disorders | 437 (15.5) |
| Neurological disorders | 350 (12.4) |
| Neurocognitive disorders | 68 (2.4) |
| Anxiety disorders | 47 (1.7) |
| Developmental disorders | 9 (0.3) |
| Other disorders | 201 (7.1) |

### Mental Health disorders

The most common mental disorder diagnosis was a psychotic disorder with peak incident presentations in the months of May and September. Mood disorders showed relatively stable trends. Substance use disorders showed occasional increases, especially in March, June, and November. Neurological disorders, other diagnoses, and neurocognitive disorders displayed moderate fluctuations throughout the year, while the pattern for anxiety and developmental disorders was difficult to decipher due to the small numbers (Fig. 2).

**Figure 2:**
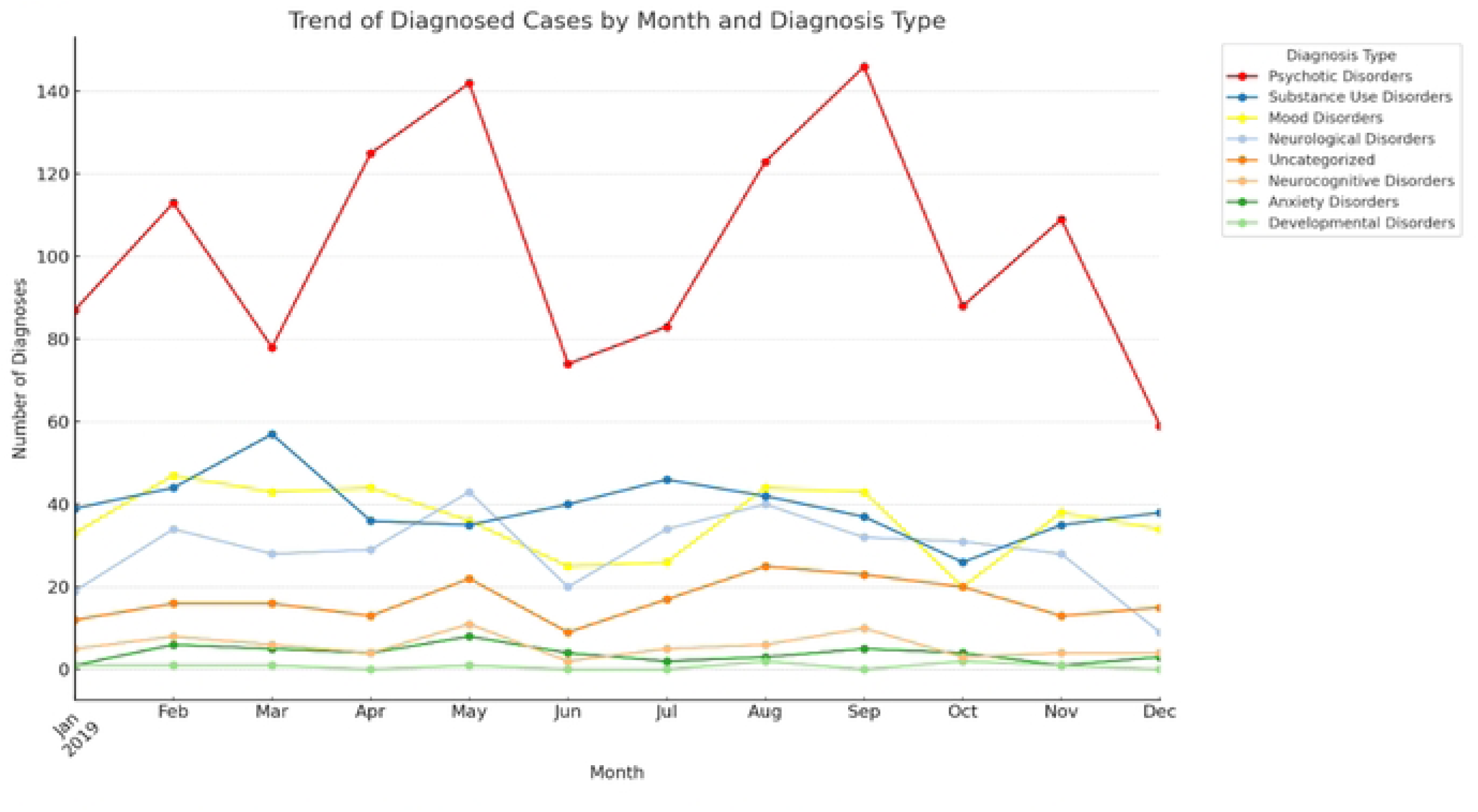
Variation of incident mental disorder admission with month of the year. Number of patients with incident presentations of different diagnoses across 2019. Mood disorders (Bipolar disorder, Depression, and Mania); Psychotic disorders (Schizophrenia, Psychosis, and Delusional disorder); Substance Use Disorders (Alcohol Use Disorder, Cannabis Use Disorder, and Substance-Induced Psychosis); Anxiety Disorders (Generalized Anxiety disorder, PTSD, Hypochondriasis and Panic Attack); Neurological Disorders (Epilepsy, Seizures, and Migraine); Neurocognitive Disorders (Dementia and Delirium); Developmental Disorders (Autism and Intellectual Disability, ADHD).

### Association of weather elements and mental disorders

The heatmap analysis shows correlations between weather variables and mental disorders (Fig. 3). Pressure shows a weak positive correlation with anxiety disorders (0.02) and a moderate positive correlation with substance use disorders (0.20), but it correlates negatively with mood disorders (-0.35). Rainfall exhibits a robust negative correlation with mood disorders (-0.40) and moderate negative correlations with anxiety (-0.35) and developmental disorders (-0.32), while having a slight positive correlation with neurocognitive disorders (0.06). Sunlight is positively correlated with mood disorders (0.33) and neurocognitive disorders (0.43), suggesting potential benefits of increased sunlight on mood and cognitive health. Humidity shows a strong negative correlation with mood disorders (-0.61) and a moderate negative correlation with developmental disorders (0.22), indicating possible adverse effects of high humidity on mood. Mean temperature has strong positive correlations with mood (0.59) and neurocognitive disorders (0.43), suggesting warmer temperatures may improve mental health outcomes. Max temperature similarly correlates positively with mood (0.55) and neurocognitive disorders (0.42), whereas min temperature exhibits a weak negative correlation with anxiety disorders (-0.32), highlighting a varied influence of temperature extremes on mental disorders.

**Figure 3:**
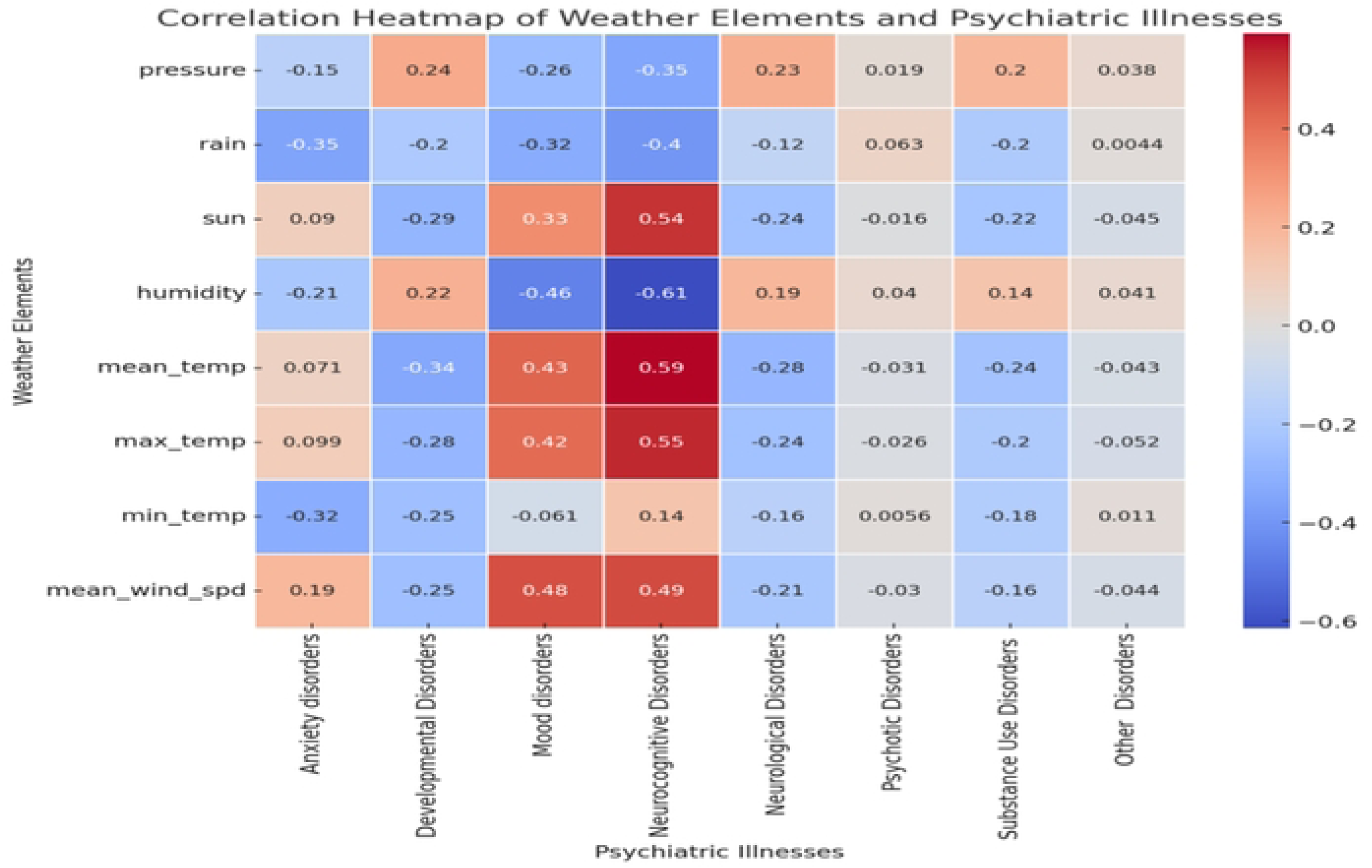
Correlations between weather elements and incident presentation for a mental disorder. Correlation coefficients between weather variables and mental illnesses, highlighting statistical relationships. Positive correlations indicate a direct association, whereas negative correlations reflect an inverse association. Data is presented for mental health outcomes including anxiety, mood, substance use, psychotic, neurocognitive, and developmental disorders.

### The role of age and sex in the association between weather elements and incident presentation for mental disorders

After controlling for age and sex, the strengths of association differed across other weather elements and incident mental disorders (Fig. 4). Each unit increase in rainfall corresponded to 5% higher odds of an incident presentation with an anxiety disorder [AOR=1.05 (95% CI: 1.01-1.22, p-value: 0.048)]. Increased rainfall was also associated with a reduced likelihood of incident presentation with a mood disorder [(OR=0.96 per mm increase, 95% CI: 0.91- 0.99, p-value: 0.034)]. Rainfall showed a significant inverse relationship with an incident presentation for a substance use disorder [OR=0.89 per mm increase (95% CI: 0.77-0.98, p-value: 0.041)]. It also showed a significant association for other disorders. An increase in sun exposure per hour per day was associated with an incident presentation with an anxiety disorder [AOR= 1.86 (95% CI: 1.56-2.32, p-value: 0.015)]. There were also significant associations between sunshine hours and incident presentation for a psychotic disorder [OR=1.12 per hour per day (95% CI: 1.09-1.21, p-value: 0.004)] and substance use disorders [OR=1.98 per hour increase (95% CI: 1.29-2.69, p-value: 0.019)]. Mean monthly temperature was associated with the incident presentation of a psychotic disorder [OR=1.67 per degree increase (95% CI: 1.06-3.32, p-value: 0.037)]. Alternatively, daily maximum temperatures were associated with incident presentations for mood disorders, psychotic disorders and substance use disorders. Other associations are shown in Table 2 below.

**Figure 4:**
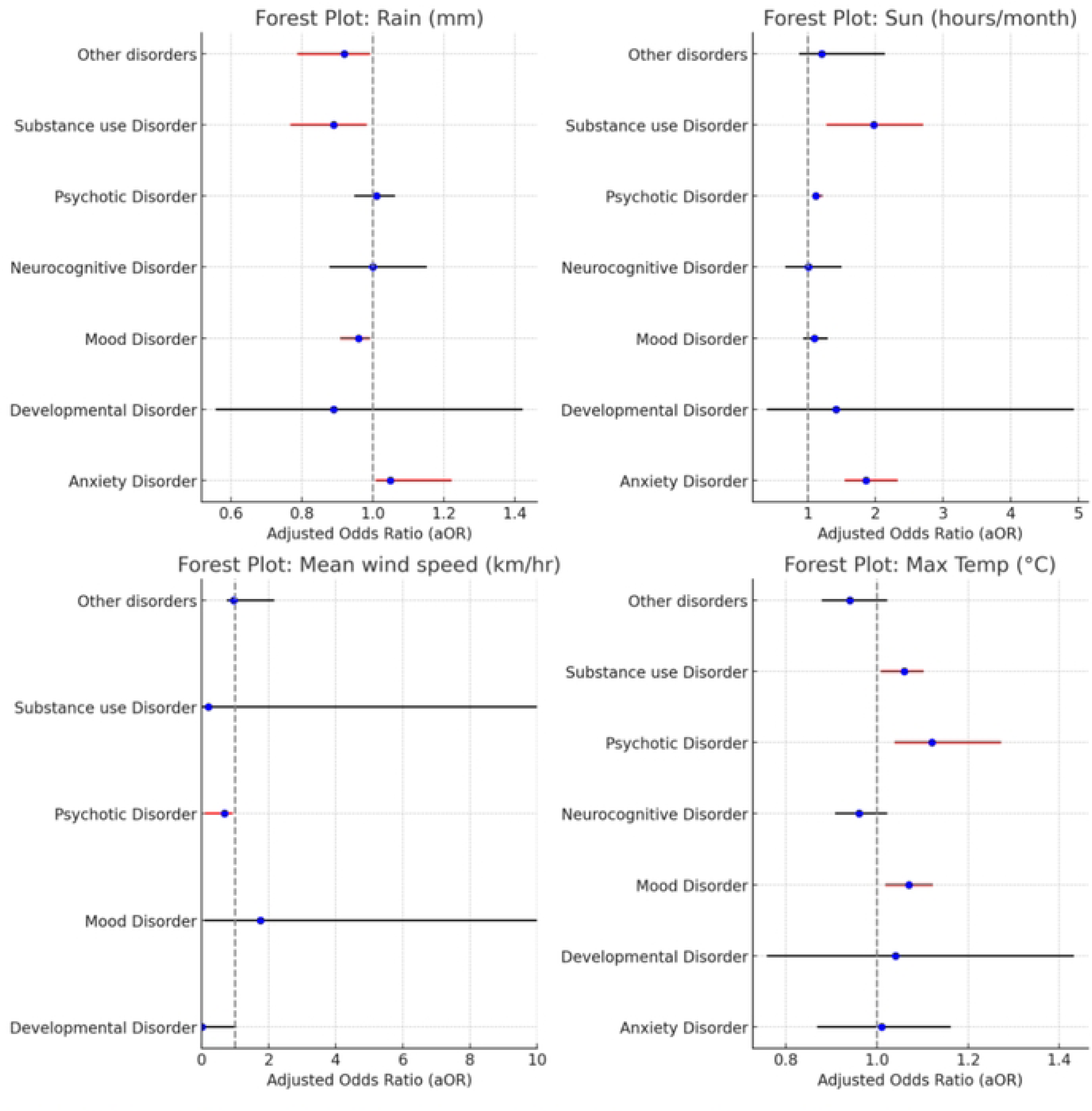
Adjusted Associations Between Weather Elements and Mental Health Outcomes. Adjusted odds ratios (aOR) illustrating the associations between specific weather elements: rainfall (A), sunshine (B), mean wind speed (C) and maximum temperature, and mental health outcomes: mood disorders, anxiety disorders, substance use disorders, and psychotic disorders, after controlling for age and sex in 2019.

**Table 2:**
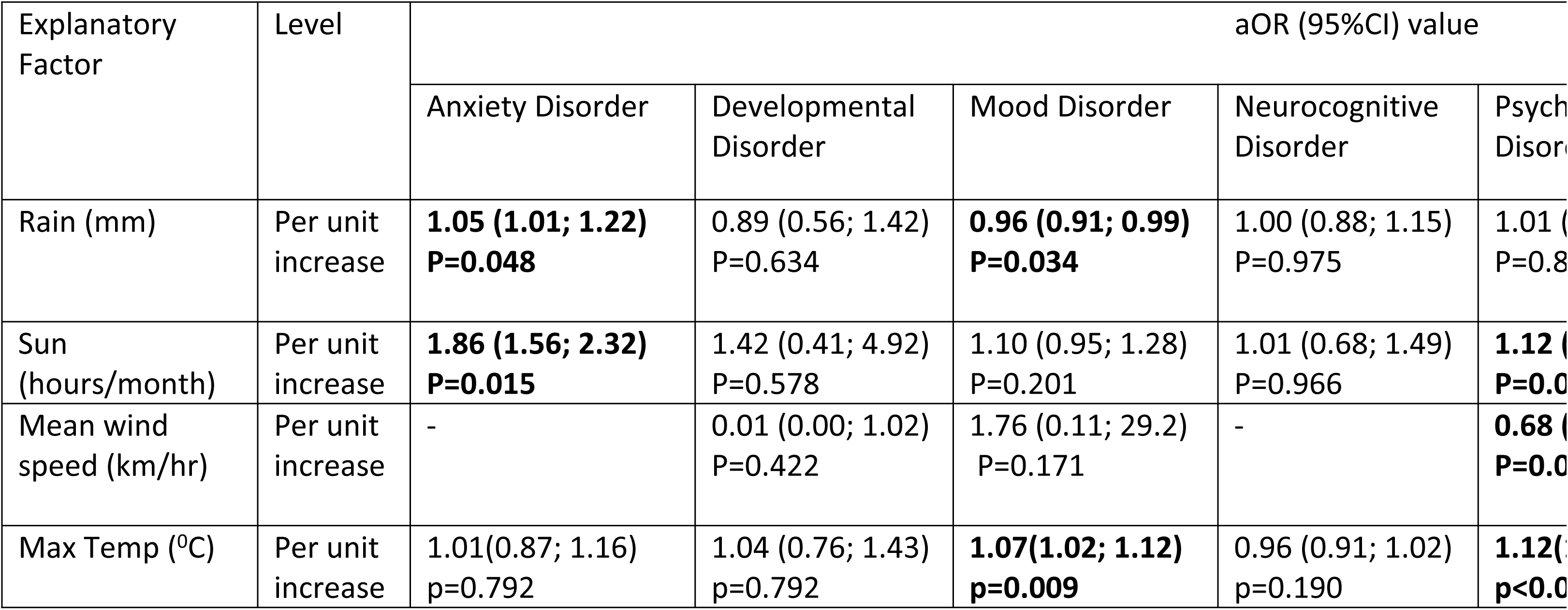
Adjusted analysis between weather factors and mental health outcomes.

## Discussion

In this study, we examined the link between specific weather elements and the incident presentation of mental disorders at Butabika National Referral Mental Hospital in Uganda. The study findings reveal significant correlations between weather elements—such as atmospheric pressure, rainfall, sunshine, temperature, and wind speed—and the incident presentation of mental disorders. Various weather elements were associated with incident presentations for psychotic and substance use disorders. Conversely, no weather elements were associated with incident presentations for neurodevelopmental and neurocognitive disorders. Age and sex modified the relationships. Rainfall was associated with incident presentations for anxiety disorders, mood disorders and substance use disorders.

The increased precipitation may have resulted in a decrease in social activity and outdoor exposure (14). This could mean fewer environmental triggers for mood disorders or less opportunity for the use of substances (15). Moreover, rain may contribute to a more subdued and calming atmosphere, reducing the likelihood of acute mental disorder episodes (16). Some of the patients with these conditions may have failed to make it to hospital due to challenging transport during a heavy downpour. For anxiety disorders, the need to stay indoors may worsen symptoms due to being kept indoors (17).

An increase in daily temperature was associated with increased odds of an incident presentation for mood, psychotic, substance and other disorders. High temperatures can lead to heat stress, dehydration, and disturbed sleep patterns, all of which are known triggers for mood swings, particularly in individuals with mood and psychotic disorders (18). For substance use disorders, higher temperatures may mean more time to indulge in substances (19). Increased periods of sunshine were associated with anxiety, psychotic and substance disorders. Sunshine increases outdoor activity and social interactions (14), which could lead to higher substance use and, consequently, a greater incidence of substance-induced psychosis or exacerbation of existing psychotic conditions. Additionally, increased sunlight exposure is known to affect serotonin levels and circadian rhythms (20), which may contribute to the onset or worsening of psychotic symptoms.

Atmospheric pressure was not associated with any mental disorders. This finding is similar to studies in South Asia (21). However, it differs from studies in temperate countries where atmospheric pressure was associated with suicide, including Finland (22). It is unclear why no weather elements were associated with an incident presentation for neurodevelopmental and neurocognitive disorders. We have previously described that caregivers are often involved in care decisions in Uganda (23). Therefore, one hypothesis for the lack of association is that elderly and young patients with these disorders are not involved in the decisions to seek care.

Our findings are an extension of previous literature that has linked weather elements with the initial presentation of various mental disorders. However, few studies have been performed in the Global South, especially in low- and middle-income countries like Uganda. Given that the Global South and especially low and middle-income countries are experiencing more significant effects of climate change (24), these findings highlight the importance of including weather variables in African mental health research.

### Future directions

Incorporating weather elements into precision psychiatry models could significantly enhance our understanding of environmental triggers in mental health. By integrating measurable weather variables—potentially monitored through wearable technologies—this approach could inform targeted interventions to mitigate climate-driven mental health risks, especially in low-resource settings where psychiatric services are limited. In the emerging field of precision psychiatry, various data points are crucial, yet few studies have considered the role of weather elements despite their measurable and pervasive nature. Seasonal affective disorder (20, 25, 26), for example, has been well-documented, but the mechanisms by which specific weather elements affect mood, psychosis, and anxiety disorders remain unclear. Thermoregulatory systems, which respond variably to weather changes, also function differently in individuals with conditions like depression and psychosis. However, the link between weather elements, thermoregulatory systems and mental disorders is still unclear. Weather elements also impact food production and availability (27), affecting mental health. This connection underscores the importance of studying weather as a factor in mental health, especially in regions where climate change increasingly threatens food security.

### Strengths and limitations

While our study offers unique insights, it assumes symptom exacerbation prompted care- seeking behaviour. Future longitudinal research could clarify additional motivators. Moreover, certain weather elements, such as atmospheric pressure, showed no association, suggesting contextual factors may influence the relevance of specific weather impacts. Despite those limitations, this study serves as a foundation for future work to explore the ecological, biological, and social pathways through which weather influences mental health, ultimately contributing to a holistic, environment-inclusive model in mental healthcare.

Further, this study introduces foundational data guiding future precision psychiatry approaches considering environmental impacts. Importantly, examining these complex interactions necessitates longitudinal studies measuring diverse variables, which could be achieved through partnerships between psychiatry, computer science and climate science. Although similar associations have been described in temperate climates, this is relatively new literature for sub-Saharan Africa (28, 29). This presents new insights into how environmental factors, including weather elements specifically and climate broadly, influence mental health incidence in low-resource settings. Future studies must incorporate additional data variables to better understand the underlying mechanisms through which weather elements impact mental disorders.

## Conclusion

This study highlights the significant relationship between weather elements and the incident presentation of mental disorders in Uganda, a low-resource setting increasingly affected by climate change. The findings underscore the importance of considering environmental factors, such as temperature, rainfall, and sunshine, in understanding mental health epidemiology and developing precision psychiatry models. While the results offer a foundation for future research, the associations identified call for further longitudinal studies to elucidate causal pathways and mechanisms. Integrating weather variables into mental health research can inform targeted interventions and public health strategies, particularly as climate change continues to pose risks to vulnerable populations. By embracing a multidisciplinary approach that includes psychiatry, climate science, and technology, we can advance towards a more holistic understanding of mental health and its environmental determinants, contributing to improved care in low-resource settings.

### Author contributions

We acknowledge the invaluable contributions of all authors to this manuscript. EKM, BTA, WS, and IM were responsible for writing the original draft, while all authors contributed to the review and editing. The conceptualization was led by EKM, WS, and IM, with investigation conducted by EKM and IM. Software contributions were handled by IM and WS, while data curation was carried out by EKM, WS, and IM. Methodology development involved FB, RK, EK, EKM, WS, IM, BBM, MB, ASS, and AK. Supervision was provided by EKM, IM, WS, SB, and BTA, and formal analysis was conducted by IM and WS. Project administration was managed by EKM, SB, and WS, with validation efforts involving all authors. Funding acquisition was led by EKM and IM, and resources were provided by EKM, WS, and IM. Finally, visualization was handled by BTA and WS.

## Acknowledgments

The authors would like to express their gratitude to the staff and administration of Butabika National Referral Mental Hospital and the Uganda National Meteorological Association for their invaluable support in providing access to the datasets utilized in this study. We also acknowledge the guidance and technical input from the Department of Psychiatry, Makerere University, and the Medical Research Council (MRC/UVRI/LSHTM) in Uganda. Special thanks to the funding agencies and all colleagues who contributed to the successful execution of this research.

## Data Availability Statement

The datasets analyzed for this study can be found in the GitHub repository for weather data https://github.com/CHAIUGA/chasa-model and the accompanying prediction web app https://github.com/CHAIUGA/chasa-webapp. The mental health datasets can be accessed via the DOI link: https://doi.org/10.1371/journal.pone.0218843.s001.

## References

1. Thompson R, Lawrance EL, Roberts LF, Grailey K, Ashrafian H, Maheswaran H, et al. Ambient temperature and mental health: a systematic review and meta-analysis. The Lancet Planetary Health. 2023;7(7):e580–e9.

2. World Health O. Mental Health and Climate change: Policy Brief: World Health Organization; 2022.

3. Clayton S, Manning C, Hodge C. Beyond Storms & Droughts: The Psychological Impacts of Climate Change. US Climate and Health Alliance. 2016.

4. Lickiewicz J, Piotrowicz K, Hughes PP, Makara-Studzińska M. Weather and Aggressive Behavior among Patients in Psychiatric Hospitals—An Exploratory Study. International Journal of Environmental Research and Public Health. 2020;17(23):9121.

5. Rajendran RK, Mohana Priya T, Jose DV, Vennira Selvi G, Poorana Senthilkumar S, Mahalakshmi SB. Stormy Minds and the Long-Term Mental Health Impact of Climate-Linked Natural Disasters. In: Samanta D, Garg M, editors. The Climate Change Crisis and Its Impact on Mental Health. Hershey, PA, USA: IGI Global; 2024. p. 1-11.

6. Wang X, Lavigne E, Ouellette-kuntz H, Chen BE. Acute impacts of extreme temperature exposure on emergency room admissions related to mental and behavior disorders in Toronto, Canada. J Affect Disord. 2014;155:154–61.

7. Li D, Zhang Y, Li X, Zhang K, Lu Y, Brown RD. Climatic and meteorological exposure and mental and behavioral health: A systematic review and meta-analysis. Sci Total Environ. 2023;892:164435.

8. Brazienė A, Venclovienė J, Vaičiulis V, Lukšienė D, Tamošiūnas A, Milvidaitė I, et al. Relationship between Depressive Symptoms and Weather Conditions. Int J Environ Res Public Health. 2022;19(9).

9. Dodds J. The psychology of climate anxiety. BJPsych Bull. 2021;45(4):222–6.

10. Jury MR. Uganda rainfall variability and prediction. Theoretical and Applied Climatology. 2018;132(3):905–19.

11. Mwesiga EK, Nakasujja N, Nakku J, Nanyonga A, Gumikiriza JL, Bangirana P, et al. One year prevalence of psychotic disorders among first treatment contact patients at the National Psychiatric Referral and Teaching Hospital in Uganda. PloS one. 2020;15(1):e0218843.

12. Association AP. Diagnostic and Statistical Manual of Mental Disorders: DSM-5. 5th ed. Arlington, VA: American Psychiatric Association; 2013.

13. Gutierrez RG. Stata. Wiley Interdisciplinary Reviews: Computational Statistics. 2010;2(6):728–33.

14. Harrison F, van Sluijs EMF, Corder K, Ekelund U, Jones A. The changing relationship between rainfall and children’s physical activity in spring and summer: a longitudinal study. International Journal of Behavioral Nutrition and Physical Activity. 2015;12(1):41.

15. Berry HL, Bowen K, Kjellstrom T. Climate change and mental health: a causal pathways framework. Int J Public Health. 2010;55(2):123–32.

16. Kandlur NR, Fernandes AC, Gerard SR, Rajiv S, Quadros S. Sensory modulation interventions for adults with mental illness: A scoping review. Hong Kong J Occup Ther. 2023;36(2):57–68.

17. Flanagan EW, Beyl RA, Fearnbach SN, Altazan AD, Martin CK, Redman LM. The Impact of COVID-19 Stay-At-Home Orders on Health Behaviors in Adults. Obesity. 2021;29(2):438–45.

18. Ebi KL, Capon A, Berry P, Broderick C, de Dear R, Havenith G, et al. Hot weather and heat extremes: health risks. The Lancet. 2021;398(10301):698–708.

19. Palamar JJ, Rutherford C, Keyes KM. Summer as a Risk Factor for Drug Initiation. Journal of General Internal Medicine. 2020;35(3):947–9.

20. Sansone RA, Sansone LA. Sunshine, serotonin, and skin: a partial explanation for seasonal patterns in psychopathology? Innov Clin Neurosci. 2013;10(7-8):20–4.

21. Ranjan J, Choudhary A, Asthana H. Atmospheric Variability and Prevalence of Common Psychiatric Disorders in South Asia: A Meta-regressive Analysis. Indian Journal of Community Health. 2019;31:289–300.

22. Hiltunen L, Ruuhela R, Ostamo A, Lönnqvist J, Suominen K, Partonen T. Atmospheric pressure and suicide attempts in Helsinki, Finland. International Journal of Biometeorology. 2012;56(6):1045–53.

23. Mwesiga EK, Nakasujja N, Nankaba L, Nakku J, Musisi S. Quality of individual and group level interventions for first-episode psychosis at the tertiary psychiatric hospital in Uganda. S Afr J Psychiatr. 2021;27:1604.

24. World Meteorological O. Africa faces disproportionate burden from climate change and adaptation costs. World Meteorological Organization. 2024.

25. Fonte A, Coutinho B. Seasonal sensitivity and psychiatric morbidity: study about seasonal affective disorder. BMC Psychiatry. 2021;21(1):317.

26. Theódórsdóttir D, Höller Y. Emotional Bias among Individuals at Risk for Seasonal Affective Disorder-An EEG Study during Remission in Summer. Brain Sci. 2023;14(1).

27. Myers SS, Smith MR, Guth S, Golden CD, Vaitla B, Mueller ND, et al. Climate Change and Global Food Systems: Potential Impacts on Food Security and Undernutrition. Annu Rev Public Health. 2017;38:259–77.

28. Smith MH. The Meteorological Factors in Mental Diseases. (Amer. Journ. Psychiat., vol. xcii, p. 131, July, 1935.) Hoverson, E. T. Journal of Mental Science. 1935;81(335):950-.

29. Tapak L, Maryanaji Z, Hamidi O, Abbasi H, Najafi-Vosough R. Investigating the effect of climatic parameters on mental disorder admissions. Int J Biometeorol. 2018;62(12):2109–18.

